# Comparison of throat swabs and sputum specimens for viral nucleic acid detection in 52 cases of novel coronavirus (SARS-Cov-2) infected pneumonia (COVID-19)

**DOI:** 10.1101/2020.02.21.20026187

**Authors:** Chenyao Lin, Jie Xiang, Mingzhe Yan, Hongze Li, Shuang Huang, Changxin Shen

## Abstract

**Background:** In December 2019, a novel coronavirus (SARS-CoV-2) infected pneumonia (COVID-19) occurred in Wuhan, China. Diagnostic test based on real-time reverse transcription polymerase chain reaction assay (qRT-PCR) was the main means of confirmation, and sample collection was mostly throat swabs, which was easy to miss the diagnosis. It is necessary to seek specimen types with higher detection efficiency and accuracy.

**Methods:** Paired specimens of throat swabs and sputum were obtained from 54 cases, and RNA was extracted and tested for 2019-nCoV (equated with SARS-CoV-2) by qRT-PCR assay.

**Results:** The positive rates of 2019-nCoV from sputum specimens and throat swabs were 76.9% and 44.2%, respectively. Sputum specimens showed a significantly higher positive rate than throat swabs in detecting viral nucleic acid using qRT-PCR assay (P=0.001).

**Conclusions:** The detection rates of 2019-nCoV from sputum specimens are significantly higher than throat swabs. We suggest that sputum would benefit for the detection of 2019-nCoV in patients who produce sputum. The results can facilitate the selection of specimens and increase the accuracy of diagnosis.

## Introduction

In December 2019, a novel coronavirus, since named severe acute respiratory syndrome coronavirus 2 (SARS-Cov-2), emerged in Wuhan, Hubei Province, China^[1]^, causing an acute febrile illness with acute respiratory distress syndrome (ARDS), which the World Health Organization (WHO) has named 2019 novel coronavirus disease (COVID-19).^[2-4]^ The type of pneumonia caused by the 2019 novel coronavirus (2019-nCoV, equated with SARS-CoV-2) is a highly infectious disease, and the WHO declared the outbreak a public health emergency of international concern on January 30, 2020.^[5]^ As of February 20, 2020, a total of 74679 clinically diagnosed and laboratory confirmed cases had been reported in China, including 2122 cases of death, and more than one thousand exported cases were reported in 26 countries spanning Asia, Europe, Oceania, and North America. The rapid global expansion and rising fatalities raised concerns about global spread.

Full-genome sequencing and phylogenic analysis indicated that 2019-nCoV was a distinct clade from the beta-coronaviruses associated with human severe acute respiratory syndrome (SARS) and Middle East respiratory syndrome (MERS).^[2]^ The 2019-nCoV is close similar to bat coronaviruses, with homology of 85-96% to a bat SARS-like coronavirus (bat-SL-CoVZC45) at the whole genome level^[6]^, and it has been postulated that bats are the primary source. However, the origin of 2019-nCoV is still under investigation, current evidence suggested that animal-to-human transmission via wild animals illegally sold in the Huanan Seafood Wholesale Market.^[7]^

A study in *The Lancet* by Huang et al^[3]^ firstly reported 41 cases of COVID-19 in which most patients had a history of exposure to Huanan Seafood Wholesale Market. The clinical manifestations of patients included fever, cough, dyspnea, myalgia, fatigue, normal or decreased leukocyte counts, and radiographic evidence of pneumonia. Organ dysfunction (such as shock, ARDS, acute cardiac injury and acute kidney injury) and death can occur in severe cases.^[3]^ Signs of infection are highly nonspecific, thus, diagnostic test based on detection of the viral sequence by real-time reverse transcription polymerase chain reaction assay (qRT-PCR) is the main means of confirmation.

Appropriate specimen selection is important for the diagnosis of respiratory viral infections.^[8]^ We collected paired specimens of throat swabs and sputum for the first time. The objective of this study was to compare the positive rates of 2019-nCoV between throat swabs and sputum specimens in the detection of viral nucleic acid by qRT-PCR. This facilitates the selection of specimens for detection of 2019-nCoV, and improve the efficiency of diagnosis for patients suspected of having COVID-19.

## Methods

### Study Design and Participants

For this retrospective, single center study, we recruited 52 patients suspected of having COVID-19 from February 7 to February 16, 2020, at Jinyintan Hospital, which located in Wuhan, Hubei Province, the endemic areas of COVID-19. Suspected patients were diagnosed according to World Health Organization interim guidance.^[9]^ All patients enrolled in this study received detection of viral nucleic acid assays (qRT-PCR) in both throat swabs and sputum specimens at the same time to make a definite diagnosis. Jinyintan Hospital is a hospital for adults (ie, aged ≥ 14 years) specialising in infectious diseases, and as well the first designated hospital for COVID-19 patients in Wuhan. The study was approved by Jinyintan Hospital Ethics Committee and oral consent was obtained from patients involved before enrolment when data were collected retrospectively.

### qRT-PCR Assay for SARS-CoV-2

Throat swabs and sputum specimens were collected for extracting 2019-nCoV RNA from patients suspected of having 2019-nCoV infection. After collection, the throat swabs were placed into a sterile test tube with 1 ml sterile saline, the sputum specimens were added equal volume of acetylcysteine (10g/L) and shaken at room temperature for 30 min to be fully liquefied, and total RNA was extracted using the nucleic acid extraction kit (QIAamp viral RNA mini kit). In brief, 40μL of cell lysates were transferred into a collection tube followed by vortex for 10 seconds. After standing at room temperature for 10 minutes, the collection tube was centrifugated at 1000 rpm/min for 5 minutes. The suspension was used for qRT-PCR assay of 2019-nCoV RNA. Then, n*19 μL mixed reagent of fluorescence PCR detection and n*1 μL RT-PCR enzyme (n was the number of reaction tubes) were mixed and vortexed for a few seconds. The above mixture of 20 μL was put into the PCR reaction tube respectively, after that the extracted sample by 5 μL was added. qRT-PCR analysis was conducted using the ABI 7500 Real-Time PCR System. The PCR parameters were 45 °C for 10 min, 95 °C for 3 min, followed by 45 cycles of 95 °C for 15 s, 58 °C for 30 s, and a single fluorescence detection point at 58°C. Two target genes, including open reading frame 1ab (ORF1ab) and nucleocapsid protein (N), were simultaneously amplified and tested during the qRT-PCR assay. The qRT-PCR assay was performed using a 2019-nCoV nucleic acid detection kit according to the manufacturer’s protocol (Shanghai ZJ Bio-Tech Co Ltd). A cycle threshold value (Ct-value) less than 37 was defined as a positive test result, and a Ct-value of 40 or more was defined as a negative test. These diagnostic criteria were based on the recommendation by the national institute for viral disease control and Prevention (China) (http://ivdc.chinacdc.cn/kyjz/202001/t20200121_211337.html). A medium load, defined as a Ct-value of 37 to less than 40, required confirmation by retesting.

### Statistical Analysis

All analyses were performed using SPSS 22.0. Continuous variables were described using mean (SD) if they are normally distributed or median (IQR) if they are not. Categorical variables were expressed as frequencies (percentages), and performed using the McNemar’s test. P<0.05 was considered statistically significant.

## Results

### Presenting Characteristics

The study population included 52 hospitalized patients suspected of having COVID-19. The average age was 57.3 years (SD, 12.5; range, 34-84 years), and 27 (51.9%) were men. All patients received qRT-PCR assays in both throat swabs and sputum specimens. (Table 1)

**Table 1.**
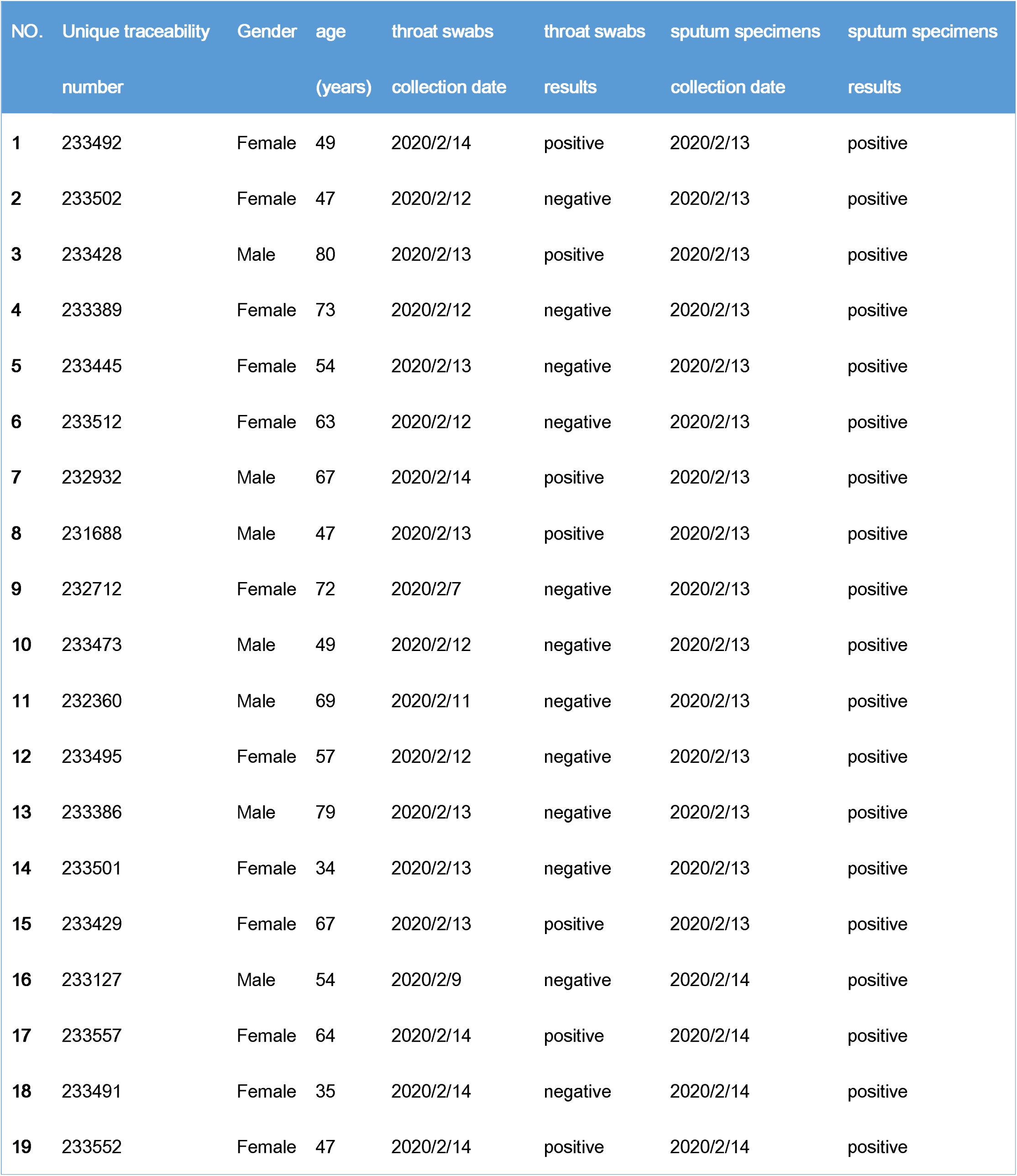
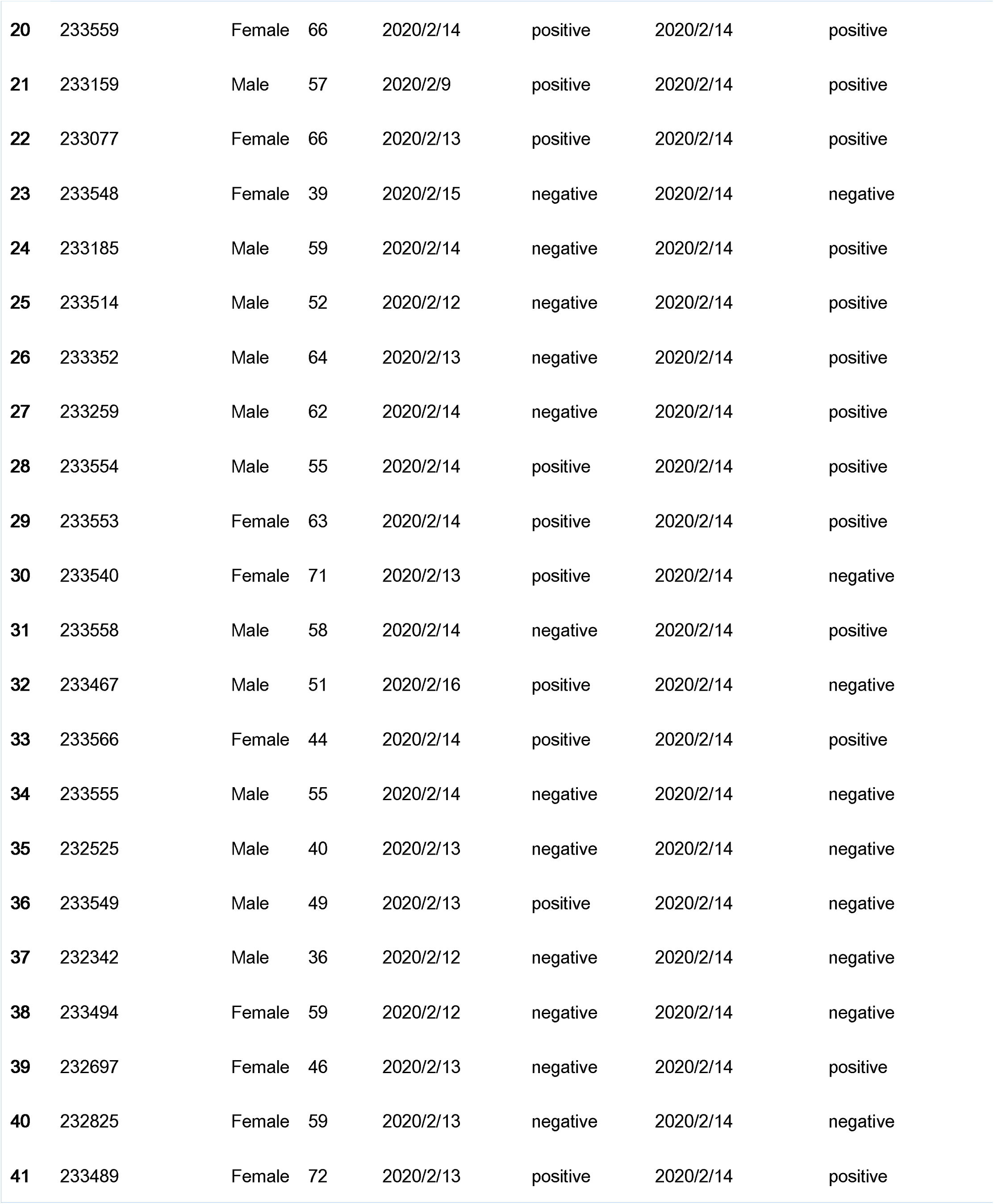
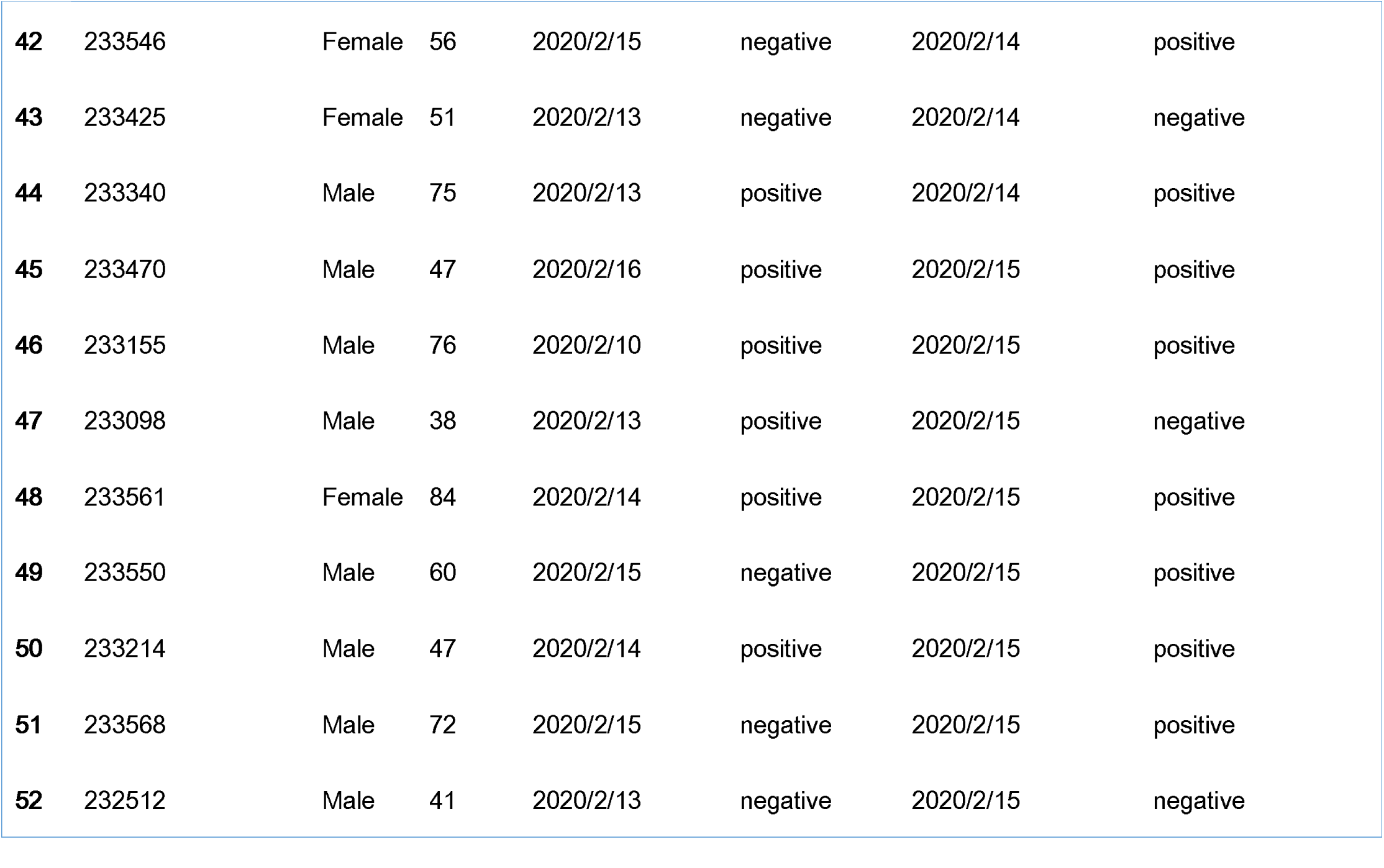
The clinical data and nucleic acid detection results (qRT-PCR) of 52 patients

### Comparison of throat swabs and sputum specimens

The viral nucleic acid by qRT-PCR showed that 23 cases (44.2%) of throat swabs were positive while 29 cases (55.8%) were negative, and 40 cases (76.9%) of sputum specimens were positive while 12 cases (23.1%) were negative (Figure 1). The positive rate of sputum specimens was almost 2-fold that of throat swabs.

**Figure 1.**
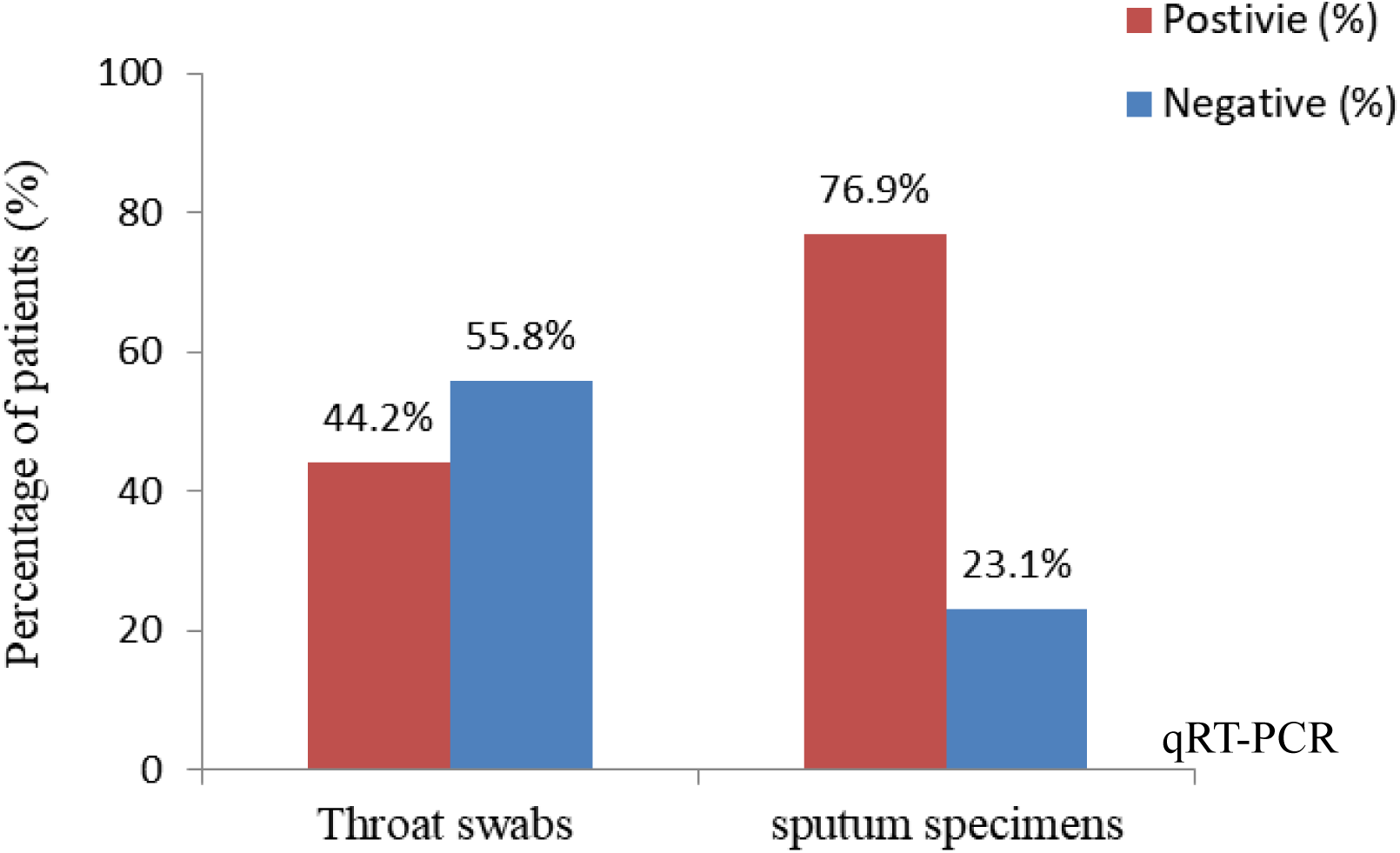
Distribution of qRT-PCR results on throat swabs and sputum specimens in patients with suspected COVID-19.

Most of the patients (51.9%) had the same results of qRT-PCR assay on throat swabs and sputum specimens, 36.5% with both positive and 15.4 % with both negative. However, quite a few patients (40.4%) showed positive sputum specimens and negative throat swabs, while only a tiny minority of patients (7.7%) showed negative sputum specimens and positive throat swabs. The findings showed that positive rates displayed a significant statistical difference (P=0.001) between throat swabs and sputum specimens. (Table 2)

**Table 2.** Comparison of qRT-PCR results between throat swabs and sputum specimens

|  | No. (%) of sputum specimens result |  |
| --- | --- | --- |
|  | Positive | Negative |
| No. (%) of throat swabs result |  |  |
| Positive | 19 (36.5%) | 4 (7.7%) |
| Negative | 21 (40.4%) | 8 (15.4%) |

*P* value = 0.001 by McNemar test.

## Discussion

Adequate specimen collection is important for the diagnosis of respiratory viral infections.^[8]^ At present, the sample collection for viral nucleic acid detection of suspected patients with COVID-19 is mostly upper respiratory tract samples (mainly throat swabs).^[10]^ The collection of throat swabs is not standardized and it is easy to miss the diagnosis. The collection process is extremely risky for medical staff. Sputum is representative of the lower respiratory tract but is rarely used for viral testing. The aim of this study was to compare the detection rates of 2019-nCoV RNA from throat swabs and sputum specimens using the qRT-PCR assay.

In our research, most of the patients were middle-aged and elderly men, which was similar to previous studies.^[3,4]^ Paired specimens of throat swabs and sputum were obtained from 52 subjects. The positive rates of 2019-nCoV from sputum specimens and throat swabs were 76.9% and 44.2%, respectively. The present study found that the overall positive rate from sputum specimens in adults was significantly higher than that from throat swabs using qRT-PCR assay. This finding was consistent with the result of a previous study that sputum showed a significantly higher positive rate than nasopharyngeal swabs in detecting respiratory viruses.^[8]^ Falsey et al^[11]^ also proved that sputum samples showed higher diagnostic yields than nose–throat swabs in adults.

The detection rate of 2019-nCoV from sputum samples was higher than that from throat swabs, which may be related to novel coronavirus’s main invasion and infection of lower respiratory tract cells, resulting in clinical manifestations such as cough and pneumonia. In addition, several studies have shown that respiratory viruses were increasingly recognized as the cause of lower respiratory tract infections.^[8]^ In these cases, specimens of the lower respiratory tract should be collected for detection of 2019-nCoV. And caution should be taken when using the negative result of viral nucleic acid from throat swabs as the criterion for the exclusion of infection and conformation of cure.

It should be noted that the high viscosity of sputum can make it difficult to extract nucleic acid. Therefore, pre-treatment of sputum samples is required, but currently no standardized pre-treatment procedure is available for virus detection. In our study, acetylcysteine was used for the digestion of sputum, which may cause substantial loss of RNA due to the washing and pipetting steps in the procedures. Therefore, thoroughly mixing and homogenization of sputum samples might be important factors to obtain reliable test results.^[8]^

This study has several limitations. First, only 52 patients with suspected COVID-19 were included, and further studies were needed to investigate the observation in a larger group of patients. Second, our study only involved throat swabs and sputum samples, and expand discussions of bronchoalveolar lavage fluid and blood samples are necessary in the future. Although sputum is more sensitive than throat swabs for 2019-nCoV detection, the use of sputum may be limited because not all patients with COVID-19 produce sputum, especially elderly patients. Therefore, if a sputum sample is available, it might be a rather reliable specimen for nucleic acid detection of 2019-nCoV.

In conclusion, the detection rates of 2019-nCoV from sputum specimens are significantly higher than those from throat swabs using the qRT-PCR assay. We suggest that sputum would benefit for the detection of novel coronavirus in patients who produce sputum. The results can facilitate the selection of specimens and increase the accuracy of diagnosis of COVID-19.

## Data Availability

The original data can be obtained in the manuscript.

## Acknowledgements

This work was supported by the Zhongnan Hospital of Wuhan University Science, Technology and Innovation Seed Fund under Grant znpy2017022.

